# Lessons Learned from Health Coverage Expansion in Massachusetts: Primary Care for Individuals with Serious Psychiatric Illness: Pre- and Post- 2006 Coverage Mandates

**DOI:** 10.1101/2023.02.01.23285355

**Authors:** Beth L. Murphy, Emily Mellen, Dominic Hodgkin, Michael T. Doonan, Bruce M. Cohen

**Author notes:** **Corresponding Author** Beth L. Murphy, M.D., Ph.D., Instructor, Harvard Medical School, Boston MA,Department of Psychiatry, McLean Hospital, 115 Mill St., Belmont, MA 02478.

## Abstract

**Purpose:** Improved access to care, particularly primary care, is a key goal of healthcare reform proposals. Individuals with severe psychiatric illness have high rates of emergency room use rather than primary/preventive care. Massachusetts implemented health care reform in 2006 intended to provide more individuals with health care insurance and, thereby, access to primary care. To our knowledge no study has assessed whether this legislation impacted barriers to primary care access among individuals with serious mental illness.

**Methods:** This study looked at effects of the 2006 Massachusetts legislation among individuals being hospitalized at a large psychiatric hospital drawing patients from throughout Eastern Massachusetts. A retrospective review of records noted whether a primary care physician was identified, along with demographic and clinical characteristics for each patient.

**Results:** Primary care affiliation was significantly lower in 2008 than 2005. Affiliation increased 9 years after legislation, though not to the levels of the year prior to the legislation. Risk ratios for PCP non-affiliation were similar whether the model controlled for demographic characteristics only; primary and drug and alcohol related diagnoses in addition to demographic characteristics; or insurance type in addition to demographic characteristics and diagnoses.

**Conclusions:** The risk of being unaffiliated with a PCP 2 years after legislative reform was nearly 20% higher than 1 year before. Our findings are concerning in that patients at this large regional psychiatry hospital may have been receiving less primary/preventive care in 2008 than in 2005, despite legislation aimed at improving primary care access for the general population.

## Introduction

There is continuing concern both about disparities in access to health care.^1^ Individuals with psychiatric disorders, among others, carry a disproportionate burden of illness and poor access to care. A 10-20 year reduction in life expectancy is seen with serious mental illness (SMI).^2-5^ Increased medical illness appears based on physiologic abnormalities, unhealthy behaviors associated with the psychiatric illness, and side effects of psychiatric medications.^4,6,7^ Inherited factors underlying psychiatric illness may also be risk factors for other medical disorders.^8-11^

Between 50-90% of individuals with SMI have chronic substantial medical co-morbidity.^12^ Further, medical co-morbidities in individuals with SMI are more costly to treat than the same illness in a psychiatrically-well group.^13,14^ Worldwide, SMI is one of the most expensive non-communicable diseases and is expected to account for one-third of total illness burden over the next two decades.^15^

In the United States, health coverage expansion legislation (HCEL) has proceeded at both state and federal levels to address access to care. One goal of HCEL is shifting patient’s primary care from emergency room visits to primary care settings, with the goal of decreasing the acuteness of patients presenting for treatment. In high-risk populations, preventive care is considered the best use of limited healthcare resources.^16^ This may be particularly true for patients with mental health and medical co-morbidities.^17^ Notably, despite the fact that the medical diseases most often seen in patients with psychiatric disorders are largely preventable or modifiable, individuals with psychiatric illness are high users of emergency services.^18-21^

Whether or not a patient has a primary care provider (PCP) is one important metric used to evaluate a patient’s access to preventive services. A robust literature details the role of improved access to PCPs in decreasing health disparities, reducing healthcare costs, and improving overall health outcomes.^22-24^ Individuals with SMI who receive primary care services have decreased morbidity and mortality^25-26^ and lacking a PCP is associated with an increased Burden of Suffering.^27^

Previous qualitative work has identified several barriers for individuals with SMI to connect with a PCP.^28^ Healthcare expansion legislation is a key mechanism through which policy-makers have aimed to address health system level barriers. However, to our knowledge no study has assessed whether HCEL legislation has helped individuals with SMI.

HCEL efforts in Massachusetts provide a unique opportunity to explore this question. In 2006, a major Health Care Reform bill was passed in Massachusetts, expanding and requiring access to insurance coverage for all state residents.^29,30.^ This initiative expanded access to state Medicaid for adult individuals with incomes of up to 133% to 150% of the Federal Poverty Level (FPL) and provided sliding scale insurance for individuals with incomes between 150-300% of FPL.^31^

We collected data on insurance and PCP affiliation from hospital admissions across three timepoints: before the 2006 Massachusetts HCEL, immediately after HCEL, and 10 years after HCEL. We assessed the effect of the 2006 Massachusetts HCEL for a number of reasons. First, Massachusetts HCEL reforms and the more recent US Patient Protection and Affordable Care Act (ACA) have key features in common: the expansion of Medicaid, subsidies of commercial insurance for low-income individuals, and both individual and business mandates.^32,33^ However, the ACA was not implemented in Massachusetts until 2014, whereas the Massachusetts HCEL was implemented in 2006, allowing for a greater opportunity to assess the long term effect of this policy.

The primary aim of the analysis was to estimate and compare risk of lacking a PCP for a population of psychiatric inpatients in 2005 and 2008, before and after implementation of HCEL legislation in Massachusetts. Secondary aims were to 1) compare risk between 2008 and 2015 to assess rebound in PCP affiliation, 2) identify demographic and clinical predictors of being without PCP affiliation.

## Methods

McLean Hospital, with 6000 annual psychiatric admissions, provides more than half the psychiatric care of the Mass General Brigham system as well as taking many referrals from other Massachusetts hospitals. It is the largest acute inpatient psychiatric care provider in Massachusetts and draws patients from all of the Eastern and much of the Western state. McLean contracts with most commercial insurances, Medicare and MassHealth/Medicaid, as well as admitting free-care/uninsured individuals.

Individuals in this study were included based on date of admission, beginning on the first day of each sample year, and proceeding chronologically until the desired N was reached. The study neither selected nor screened-out people with specific characteristics, such as ethnicity, religion, sex, or sexual orientation. Individuals over 65 were excluded, as non-disability Medicare coverage would not likely be affected by HCEL implementation. Only data from first admission in any year was included.

The year prior to introduction of HCEL, 2005, was the ‘control’ year. As 2007 was a transitional year, 2008 was chosen for assessing early effects of HCEL. To explore long -term consequences of HCEL, data were collected from 2015.

Individuals were grouped by diagnostic category, with priority defined as the diagnosis likely to have the greatest impact on level of functioning. Specifically, priority was defined as: psychotic disorders> bipolar disorders> other mood disorders (predominantly unipolar depression and anxiety). For example, an individual with schizoaffective disorder was coded as ‘psychotic disorder’. Individuals with substance abuse/dependence and an axis I disorder were counted under substance abuse/dependence, unless axis I diagnosis was a psychotic disorder.

Insurances were grouped as: Commercial, Medicare, Medicaid/Free Care, which were mutually exclusive. (In Massachusetts, Free Care is managed by the same teams that coordinate care for Medicaid-covered individuals.)

De-identified information was entered into a computerized database on a Microsoft Access Platform.

### Sample

Records were reviewed from a total of 2532 admissions: the first 1035 admissions from 2005, the first 1151 admissions from 2008, and the first 346 admissions from 2015. The sample size from 2015 was smaller because data were collected for a secondary analysis, to determine if rates of PCP affiliation would change over time. Eliminating second admissions in the same year left 733 2005 admissions, 842 2008 admissions, and 318 2015 admissions. Because the new legislation only applied to Massachusetts residents, we limited the analytic sample to state residents. This left 678 2005 admissions, 786 2008 admissions, and 250 2015 admissions.

### Statistical Approach

Demographic and clinical characteristics were summarized using means and standard deviations (age) and frequencies and percentages (categorical variables). Characteristics were compared between years using linear regression, binary logistic regression, and multinomial logistic regression, as appropriate. To address primary aims of estimating the risk of being unaffiliated with a PCP and comparing risk before and after the introduction of HCEL, percentage of patients unaffiliated with a PCP was calculated for each year, and corresponding 95% confidence intervals (CIs) were calculated using Wilson’s method.^34^ Risk regression compared relative risk of being unaffiliated with a PCP between years, controlling for demographic and clinical characteristics of patients and shifts in type of insurance coverage. To assess changes in demographic, clinical, and insurance profiles of patients between calendar years, control covariates were added to our models. The first stage model included demographic covariates only (age, sex, and ethnicity), the second stage model included demographic and diagnostic covariates (primary diagnosis and presence of drug or alcohol abuse or dependence), and the third stage model included demographic and clinical covariates and insurance type (Medicaid, Medicare, or commercial) along with demographic and diagnostic covariates. The relative risk estimate from the second stage model was expected to best quantify the impact of healthcare legislation because changes in type of insurance, but not changes in the demographic and clinical profile of the patients between admission years, were likely to have been substantially impacted by HCEL. The third stage model assessed the degree to which changes between years could be explained by shifts in type of insurance coverage.

Relative risk estimates for demographic and clinical covariates in these models addressed the secondary aim of identifying demographic and clinical predictors of PCP affiliation. To assess differential impact of HCEL by psychiatric diagnosis, an interaction between admission year and psychiatric diagnosis was added to the second stage model. To assess the impact of confounding by changes in the patient population between admission years not accounted for in primary analysis, a secondary analysis examined within-patient changes in PCP affiliation for the subset of patients admitted in both 2005 and 2008 (97 individuals) using McNemar’s test.^35^

Regression models were fit using PROC GENMOD in SAS (version 9.4, SAS Institute, Cary, NC), with the method of generalized estimating equations to account for clustering of admissions from the same individuals. The modified Poisson approach for binary data was used to perform relative risk regression.^36^ Admissions with missing data were omitted from corresponding statistical models. Statistical significance required test-wise two-tailed p<0.05.

### Compliance with Ethical Standards

This work was approved by the Partners Institutional Review Board (protocol number 2011P001111). The authors have no financial disclosures to report.

### Data Availability

Data for this investigation was gathered from de-identified electronic medical records. Due to the sensitive and confidential nature of EMRs, these data are not available publicly.

## Results

Characteristics of the sample by year of admission are in Table 1. Gender ratio was near 50% all three years. Mean age, which was between 35 and 40 for the three years, was similar between 2005 and 2008 but younger in 2015 than 2008. Between 2005 and 2008 there was a shift away from Medicare and toward commercial insurance. Across the three years there were fewer admissions for diagnoses with psychosis and more non-bipolar admissions without psychosis. Between 2008 and 2015 there was an increase in the percentage of patients admitted with alcohol or drug abuse or dependence.

**Table 1:** Characteristics of the Sample by Year of Admission.

|  | 2005 | 2008 | 2015 | p<sup>1</sup> | p<sup>1</sup> |
| --- | --- | --- | --- | --- | --- |
|  | N=678 | N=786 | N=250 | 2008 vs.<br>2005 | 2015 vs.<br>2008 |
| Age, Mean (SD) | 38.6<br>(12.5) | 38.8<br>(12.7) | 35.6<br>(13.2) | 0.76 | <0.001 |
| Female, N(%) | 321<br>(47%) | 382<br>(49%) | 132<br>(53%) | 0.61 | 0.25 |
| Non-Hispanic White, N(%)<sup>2</sup> | 582<br>(87%) | 672<br>(86%) | 207<br>(86%) | 0.78 | 0.88 |
| Insurance Type<sup>3</sup> |  |  |  | 0.001 | 0.12 |
| Medicare | 262<br>(39%) | 248<br>(32%) | 60<br>(25%) |  |  |
| Commercial | 318<br>(47%) | 440<br>(56%) | 152<br>(63%) |  |  |
| Medicaid/ Free care | 95<br>(14%) | 95<br>(12%) | 30<br>(12%) |  |  |
| Primary Diagnosis<sup>4</sup> |  |  |  | <0.001 | <0.001 |
| Psychosis | 378<br>(56%) | 352<br>(45%) | 50<br>(20%) |  |  |
| Bipolar Disorder Without<br>Psychosis | 156<br>(23%) | 167<br>(21%) | 37<br>(15%) |  |  |
| Non-Bipolar Mood Disorder<br>Without Psychosis | 120<br>(18%) | 236<br>(30%) | 125<br>(50%) |  |  |
| Other or No Axis I Diagnosis | 21<br>(3%) | 28<br>(4%) | 37<br>(15%) |  |  |
| Alcohol or Drug Abuse or<br>Dependence<sup>4</sup> | 229<br>(34%) | 274<br>(35%) | 129<br>(52%) | 0.66 | <0.001 |

1. Characteristics were compared between years using linear regression (age), binary logistic regression (white and alcohol or drug abuse or dependence), and multinomial logistic regression (insurance type and primary diagnosis) with robust standard errors. 2. Ethnicity was missing for 7 of the 2005 patients, 7 of the 2008 patients, and 9 of the 2015 patients. 3. Insurance type was missing for 3 of the 2005 patients, 3 of the 2008 patients, and 8 of the 2015 patients. 4. Diagnostic information was missing for 3 of the 2005 patients, 3 of the 2008 patients, and 1 2015 patient.

The proportion of patients unaffiliated with a PCP was 53.4% (95% CI 49.6%: 57.1%) among those admitted in 2005, 61.8% (95% CI 58.4%: 65.2%) in 2008, and 56.4% (95% CI 50.2%: 62.4%) in 2015. Adjusted risk ratios comparing being unaffiliated with a PCP in 2008 versus 2005, and 2015 versus 2008, are in Table 2. Non-affiliation was significantly higher in 2008 than 2005 (nearly 20% increased, p<0.001, all three models). Estimated risk decreased in 2015, though not to the level estimated for 2005, and not statistically different from 2008. Relative risk estimates were similar after controlling for demographic characteristics only; primary and drug and alcohol related diagnoses in addition to demographic characteristics; and insurance type in addition to demographic characteristics and diagnoses. (That is, relative risk estimates from stage one, two, and three models were similar). Two patients admitted in 2005, five patients admitted in 2008, and no patients admitted in 2015 lacked information about a PCP recorded in their admission records and were excluded from analyses.

**Table 2:** Risk ratios (RRs) for being unaffiliated with a PCP, associated 95% confidence intervals, and results of significance tests.

|  | RR<sub>1</sub> (95%<br>CI) | p<sub>1</sub> | RR<sub>2</sub> (95%<br>CI) | p<sub>2</sub> | RR<sub>3</sub> (95%<br>CI) | p<sub>3</sub> |
| --- | --- | --- | --- | --- | --- | --- |
|  | N=1684 |  | N=1677 |  | N=1664 |  |
| Year of admission |  |  |  |  |  |  |
| 2008 vs. 2005 | 1.17 (1.07,<br>1.28) | <0.001 | 1.18 (1.08,<br>1.29) | <0.001 | 1.19 (1.09,<br>1.30) | <0.001 |
| 2015 vs. 2008 | 0.91 (0.81,<br>1.03) | 0.13 | 0.94 (0.83,<br>1.07) | 0.33 | 0.92 (0.81,<br>1.04) | 0.19 |
| Age, in decades | 0.96 (0.93,<br>0.99) | 0.02 | 0.96 (0.93,<br>0.99) | 0.02 | 0.95 (0.92,<br>0.99) | 0.008 |
| Female | 0.87 (0.80,<br>0.94) | <0.001 | 0.86 (0.79,<br>0.94) | <0.001 | 0.86 (0.79,<br>0.93) | <0.001 |
| Non-white | 0.96 (0.85,<br>1.08) | 0.51 | 0.96 (0.85,<br>1.08) | 0.47 | 0.96 (0.85,<br>1.08) | 0.48 |
| Primary diagnosis |  |  |  | 0.10 |  | 0.14 |
| No psychosis or<br>BPD |  |  | --- |  | --- |  |
| BPD without<br>psychosis |  |  | 1.13 (1.00,<br>1.26) |  | 1.12 (1.00,<br>1.26) |  |
| Psychosis |  |  | 1.09 (0.99,<br>1.21) |  | 1.08 (0.97,<br>1.20) |  |
| Drug or alcohol<br>diagnosis |  |  | 0.97 (0.89,<br>1.06) | 0.55 | 0.97 (0.89,<br>1.06) | 0.54 |
| Insurance type |  |  |  |  |  | 0.21 |
| Medicaid/free care |  |  |  |  | --- |  |
| Medicare |  |  |  |  | 1.10 (0.96,<br>1.26) |  |
| Commercial |  |  |  |  | 1.02 (0.89,<br>1.16) |  |

Risk ratios, confidence intervals, and p-values were obtained from risk regression models controlling for demographic predictors (models 1-3), diagnosis (models 2-3), and insurance type (model 3 only). Because admissions with missing data were omitted, the number of admissions contributing data (N) differed between models. Primary diagnosis categories and insurance type are mutually exclusive.

Results from the secondary analysis examining changes in PCP affiliation among patients with admissions in both 2005 and 2008 supported the results from the primary analysis. Among the 93 patients who had admissions and non-missing PCP affiliation in both 2005 and 2008, 38 (41%) changed PCP affiliation status. These 38 were significantly more likely to change from affiliation to non-affiliation (n=28) than to change from non-affiliation to affiliation (n=10, S=8.53, p=0.004 McNemar’s test).^35^

Across all years, risk of being unaffiliated with a PCP was lower for older patients and females. There was no significant association with primary diagnosis, diagnosis of drug or alcohol abuse or dependence, or type of insurance. Association with diagnosis did not differ significantly between 2005 and 2008 (χ^2^(2) =1.25, p=0.53)

## Discussion

Following HCEL implementation in Massachusetts, a greater percentage of psychiatric inpatients in our sample were covered by commercial insurance. Despite the changes in insurance, the risk of being unaffiliated with a PCP in 2008, following HCEL, was nearly 20% higher than it was in 2005. Although unaffiliation decreased in 2015, it did not reach the level observed for 2005, and there was no statistically significant difference from 2008.

These results are surprising, considering that the Massachusetts 2006 HCEL program has been judged successful in many ways: implementation has been associated with an increase in health insurance among residents, a decrease in individuals reporting they are delaying or omitting health care due to cost, a decrease in low severity emergency room visits, and an increase in the proportion of residents having a PCP.^29,31^ These benefits were also noted in certain socioeconomically disadvantaged groups, such as Hispanic and lower income individuals.^32^ That said, some investigations have noted no appreciable decrease in unreimbursed costs for the uninsured in government programs and safety-net hospital settings, suggesting that the success of HCEL may be more limited in contexts with larger percentages of uninsured patients.^37^ Among our sample of psychiatric inpatients, the results suggest that there are barriers to PCP access beyond the availability of insurance.

One possible explanation for these findings is that the supply of PCPs was inadequate to cover the increased number of individuals seeking a practice affiliation after implementation of HCEL. Some disadvantaged groups reported an increase in primary care access after the 2006 HCEL.^32^ Individuals with SMI, on average, did not, suggesting that having an SMI may impair an individual in effectively searching for and achieving access to a primary care physician.

This study has limitations. First, the study sample was recruited from a single psychiatric hospital and data regarding PCP affiliation is not available on a statewide basis. Second, the Boston area neighborhoods that include a higher percentage of specific racial/ethnic populations may be less likely to refer to McLean hospital than to other units. However, McLean inpatients are form a diverse population, predominantly across Eastern Massachusetts, where most of the state population lives, and their demographics were stable over time, suggesting that changes seen were associated with the implementation of HCEL.

The federal Affordable Care Act (ACA) was passed in 2010 and implemented in Massachusetts in 2014, replacing the 2006 HCEL. Given that the ACA had only been implemented for a few months at the time of our 2015 sample, the impact of the 2014 implementation of the ACA on our sample was likely negligible. By comparison, given the effects of the HCEL on PCP affiliation for those with SMI in Massachusetts, impacts of the ACA on similar individuals elsewhere might have been considerable.

## Conclusions

This study found an increase in commercial insurance coverage but a decline in PCP affiliation following the introduction of HCEL in Massachusetts among individuals receiving treatment for serious psychiatric illnesses. This finding is both striking and concerning. Ensuring that individuals with SMI receive effective preventive care will be critical for healthcare coverage and preventive care utilization plans to be successful in reaching their goals. Our findings suggest a need for similar research into the effects of ACA on psychiatric patients’ access to primary care, particularly in states that expanded Medicaid. In addition, in Massachusetts, additional research will be required to clarify group and diagnosis-specific barriers in order to consider effective interventions.

## Data Availability

All data produced in the present study are available upon reasonable request to the authors

## Notes

### Competing Interest Statement

The authors have declared no competing interest.

### Funding Statement

This study did not receive any funding

### Author Declarations

McLean Hospital is part of Partners Healthcare (now called Mass General Brigham) IRB protocol #2001P001111

## References

1. Branning G, Vater M. Healthcare spending: plenty of blame to go around.Am Health Drug Benefits. 2016;9(8):445–447.

2. Chang CK, Hayes RD, Perera G, Broadbent MT, Fernandes AC, Lee WE, Hotopf M, Stewart R. Life expectancy at birth for people with serious mental illness and other major disorders from a secondary mental health care case register in London. PLoS One. 2011;6(5):e19590. doi:10.1371/journal.pone.0019590

3. Hayes RD, Chang CK, Fernandes AC, Begum A, To D, Broadbent M, Hotopf M, Stewart R. Functional status and all-cause mortality in serious mental illness. PLoS One. 2012;7(9):e44613. doi:10.1371/journal.pone.0044613

4. Viron MJ, Stern TA. The impact of serious mental illness on health and healthcare. Psychosomatics. 2010;51(6):458–465. doi:10.1176/appi.psy.51.6.458

5. Felker B, Yazel JJ, Short D. Mortality and medical comorbidity among psychiatric patients: a review. Psychiatr Serv. 1996;47(12):1356–1363. doi:10.1176/ps.47.12.1356

6. Newcomer JW. Antipsychotic medications: metabolic and cardiovascular risk. J Clin Psychiatry. 2007;68 Suppl 4:8–13.

7. Fagiolini A, Goracci A. The effects of undertreated chronic medical illnesses in patients with severe mental disorders. J Clin Psychiatry. 2009;70 Suppl 3:22–29. doi:10.4088/JCP.7075su1c.04

8. Patten SB, Williams JV, Lavorato DH, Modgill G, Jetté N, Eliasziw M. Major depression as a risk factor for chronic disease incidence: longitudinal analyses in a general population cohort. Gen Hosp Psychiatry. 2008;30(5):407–413. doi:10.1016/j.genhosppsych.2008.05.001

9. Knol MJ, Twisk JW, Beekman AT, Heine RJ, Snoek FJ, Pouwer F. Depression as a risk factor for the onset of type 2 diabetes mellitus. A meta-analysis. Diabetologia. 2006;49(5):837–845. doi:10.1007/s00125-006-0159-x

10. Chwastiak LA, Rosenheck RA, Desai R, Kazis LE. Association of psychiatric illness and all-cause mortality in the National Department of Veterans Affairs Health Care System. Psychosom Med. 2010;72(8):817–822. doi:10.1097/PSY.0b013e3181eb33e9

11. Kilbourne AM, Morden NE, Austin K, et al. Excess heart-disease-related mortality in a national study of patients with mental disorders: identifying modifiable risk factors. Gen Hosp Psychiatry. 2009;31(6):555–563. doi:10.1016/j.genhosppsych.2009.07.008

12. Almeida OP, Pirkis J, Kerse N, Sim M, Flicker L, Snowdon J, Draper B, Byrne G, Lautenschlager NT, Stocks N, Alfonso H, Pfaff JJ. Socioeconomic disadvantage increases risk of prevalent and persistent depression in later life. J Affect Disord. 2012;138(3):322–331. doi:10.1016/j.jad.2012.01.021

13. Hutter N, Schnurr A, Baumeister H. Healthcare costs in patients with diabetes mellitus and comorbid mental disorders--a systematic review. Diabetologia. 2010;53(12):2470–2479. doi:10.1007/s00125-010-1873-y

14. Hutter N, Knecht A, Baumeister H. Health care costs in persons with asthma and comorbid mental disorders: a systematic review. Gen Hosp Psychiatry. 2011;33(5):443–453. doi:10.1016/j.genhosppsych.2011.06.013

15. Insel TR, Collins PY, Hyman SE. Darkness Invisible: The hidden global costs of mental illness. Foreign Affairs. 2015;94(1):127–135. http://www.jstor.org/stable/24483225

16. Cohen JT, Neumann PJ, Weinstein MC. Does preventive care save money? Health economics and the presidential candidates. N Engl J Med. 2008;358(7):661–663. doi:10.1056/NEJMp0708558

17. Goodell S, Druss BG, Walker ER. Mental disorders and medical comorbidity. Princeton, NJ: The Robert Wood Johnson Foundation. The Synthesis Project. 2011. policy brief No. 21. https://www.rwjf.org/en/library/research/2011/02/mental-disorders-and-medical-comorbidity.html

18. Smith S, Yeomans D, Bushe CJ, Eriksson C, Harrison T, Holmes R, Mynors-Wallis L, Oatway H, Sullivan G. A well-being programme in severe mental illness. Baseline findings in a UK cohort. Int J Clin Pract. 2007;61(12):1971–1978. doi:10.1111/j.1742-1241.2007.01605.x

19. Lawrence D, Coghlan R. Health inequalities and the health needs of people with mental illness. N S W Public Health Bull. 2002;13(7):155–158. doi:10.1071/nb02063

20. Hackman AL, Goldberg RW, Brown CH, Fang LJ, Dickerson FB, Wohlheiter K, Medoff DR, Kreyenbuhl JA, Dixon L. Use of emergency department services for somatic reasons by people with serious mental illness. Psychiatr Serv. 2006;57(4):563–566. doi:10.1176/ps.2006.57.4.563

21. Niedzwiecki MJ, Sharma PJ, Kanzaria HK, McConville S, Hsia RY. Factors associated with emergency department use by patients with and without mental health diagnoses. JAMA Netw Open. 2018;1(6):e183528. doi:10.1001/jamanetworkopen.2018.3528

22. Starfield B, Shi L, Macinko J. Contribution of primary care to health systems and health. Milbank Q. 2005;83(3):457–502. doi:10.1111/j.1468-0009.2005.00409.x

23. Levine S, Malone E, Lekiachvili A, Briss P. Health Care Industry Insights: Why the use of preventive services Is still low. Prev Chronic Dis. 2019;16:E30. Published 2019 Mar 14. doi:10.5888/pcd16.180625

24. Shi L. The impact of primary care: a focused review. Scientifica (Cairo). 2012;2012:432892. doi:10.6064/2012/432892

25. US Dept of Health and Human Services. Office of Disease Prevention and Health Promotion. Clinical Preventative Services. https://health.gov/healthypeople/objectives-and-data/leading-health-indicators

26. Grudniewicz A, Peckham A, Rudoler D, Lavergne MR, Ashcroft R, Corace K, Kaluzienski M, Kaoser R, Langford L, McCracken R, Norris WC, O′Riordan A, Patrick K, Peterson S, Randall E, Rayner J, Schütz CG, Sunderji N, Thai H, Kurdyak P. Primary care for individuals with serious mental illness (PriSMI): protocol for a convergent mixed methods study. BMJ Open 2022;12:e065084. doi:10.1136/bmjopen-2022-065084

27. Olsen CG, Boltri JM, Amerine J, Clasen ME. Lacking a primary care physician Is associated with increased suffering in patients with severe mental illness. J Prim Prev. 2017;38(6):583–596. doi:10.1007/s10935-017-0490-7

28. Ross LE, Vigod S, Wishart J, Waese M, Spence JD, Oliver J, Chambers J, Anderson S, Shields R. Barriers and facilitators to primary care for people with mental health and/or substance use issues: a qualitative study. BMC Fam Pract. 2015;16:135. doi:10.1186/s12875-015-0353-3

29. Doonan MT, Tull KR. Health care reform in Massachusetts: implementation of coverage expansions and a health insurance mandate. Milbank Q. 2010;88(1):54–80. doi:10.1111/j.1468-0009.2010.00589.x

30. Kleinpeter MA. The Massachusetts health insurance law: providing health insurance to all. J Natl Med Assoc. 2006;98(11):1867–1873.

31. Kaiser Family Foundation (2012) Massachusetts Health Care Reform: Six Years Later. Focus on Health Reform. Publication 8311, (May, 2012) http://kff.org/health-costs/issue-brief/massachusetts-health-care-reform-six-years-later/

32. Kolstad JT, Kowalski AE. The impact of health care reform on hospital and preventive care: evidence from Massachusetts. J Public Econ. 2012;96(11-12):909–929. doi:10.1016/j.jpubeco.2012.07.003

33. Short-term effects of health care coverage legislation-Massachusetts 2008. CDC: Centers for Disease Control and Prevention. Morbidity and Mortality Weekly Report, March 12, 2010; 59(9), 262–267. https://www.cdc.gov/mmwr/preview/mmwrhtml/mm5909a3.htm

34. Wilson EB. Probable inference, the law of succession, and statistical inference. J Am Stat Assoc. 1927;22:209–212. doi:10.2307/2276774

35. Pembury Smith MQR., Ruxton GD. Effective use of the McNemar test. Behav Ecol Sociobiol 74, 133(2020). doi:10.1007/s00265-020-02916-y

36. Zou G. A modified poisson regression approach to prospective studies with binary data. Am J Epidemiol. 2004;159(7):702–706. doi:10.1093/aje/kwh090

37. Bazzoli GJ, Clement JP. The experiences of Massachusetts hospitals as statewide health insurance reform was implemented. J Health Care Poor Underserved. 2014;25(1 Suppl):63–78. doi:10.1353/hpu.2014.0073

